# Intraglottal aerodynamic pressure and energy transfer in a self-oscillating synthetic model of the vocal folds

**DOI:** 10.1101/2020.11.20.20235911

**Authors:** Mohsen Motie-Shirazi, Matías Zañartu, Sean D. Peterson, Byron D. Erath

**Affiliations:** Department of Mechanical and Aeronautical Engineering, Clarkson University, Potsdam, NY, USA; Department of Electronic Engineering, Universidad Técnica Federico Santa María, Valparaíso, Chile; Department of Mechanical and Mechatronics Engineering, University of Waterloo, Waterloo, ON, CA

**Keywords:** Intraglottal aerodynamics, Aerodynamic pressure, Aerodynamic energy, Synthetic vocal fold model

## Abstract

Self-sustained oscillations of the vocal folds during phonation are the result of the energy exchange between the airflow and the vocal fold tissue. Understanding this mechanism requires accurate investigation of the aerodynamic pressures acting on the vocal fold surface during oscillation. A self-oscillating silicone vocal fold model was used in a hemilaryngeal flow facility to measure the time-varying pressure distribution along the inferior-superior length of the vocal fold with a spatial resolution of 0.254 mm, and at four discrete locations in the anterior-posterior direction. It was found that the intraglottal pressures during the opening and closing phases of the vocal fold are highly dependent on three-dimensional and unsteady flow behaviors. The measured aerodynamic pressures and estimates of the medial surface velocity were used to compute the intraglottal energy transfer from the airflow to the vocal folds. The energy was greatest at the anterior-posterior midline, and decreased significantly toward the anterior/posterior endpoints. The net energy transfer over an oscillation cycle was positive, consistent with the theory of energy exchange during phonation. The findings provide insight into the dynamics of the vocal fold oscillation and the potential causes of some vocal fold disorders.

## I. INTRODUCTION

Self-sustained oscillations of the vocal folds (VFs) are the result of coupling between the aerodynamic energy that is transferred to the VFs by the air being forced between them, the biomechanical structure of the VFs, and the acoustic loading of the vocal tract^1^. The rate of energy transfer from the fluid to the VF tissue is correlated with the pressure and velocity fields within the glottis (i.e., the opening between the VFs)^2,3^. Because the glottis forms a time-varying orifice during phonation, high-fidelity temporal and spatial resolution of the intraglottal pressure and velocity is needed to elucidate the physics of VF oscillation^4,5^.

Glottal velocity fields have been widely studied using static and dynamic VF models^6–16^, excised larynges^17–20^, and computational approaches^21–29^, with a review of the literature found in Mittal et al.^30^. Surprisingly, there is little work that has connected the observed fluid flow phenomena with the resultant pressure loading at sufficient spatial and temporal resolutions to provide direct insight into the energy exchange process. Initial work utilized static VF models in steady flow^31–35^, identifying that the glottal pressure reached a minimum for divergent orientations. Subsequent work investigated the influence of features such as inferior/superior VF angles^35–37^ and inlet and exit VF radii^38–40^ on the pressure field. While providing significant spatial resolution of the VF surface pressure, the use of static geometries in steady flow, commonly justified by invoking the quasi-steady assumption^41^, neglects temporal dependencies in the flow. Subsequent work has shown the quasi-steady assumption overlooks key fluid dynamics, particularly during the opening and closing phases when flow accelerations are very high^15,42–44^.

During the closing phase of a modal phonatory cycle, the glottis forms a divergent channel. The resultant adverse pressure gradient gives rise to complex flow behavior, including variations in the flow separation point^45^, formation and propagation of vortices within the glottis^20,46–48^, and flow asymmetries. This phenomenon has been observed using particle image velocimetry (PIV) in studies with static^11^, dynamically driven^14^, and self-sustained oscillating^49^ models of the VFs as well as in an excised canine larynx^50^. The resultant pressure loading has been investigated by exploring a theoretical solution for the asymmetric pressure loading^51–53^. While this empirical approach was derived from unsteady, driven vocal-fold oscillations, the model implementation relied upon a quasi-steady assumption, and further neglected three-dimensional effects and supraglottal geometry (i.e., the ventricular folds), which have subsequently been shown to be important in determining the development of flow asymmetries^54,55^.

Finally, despite the fairly common use of two-dimensional VF model geometries that are extruded in the anterior-posterior direction, the three-dimensionality of the VFs has been shown to have a significant influence on VF surface pressure loadings^56^, exhibiting a lower pressure drop than that observed for two-dimensional models, and giving rise to anterior-posterior variations in the pressure loading. Unfortunately, these effects have largely been investigated in static models with steady flow. A notable exception measured the unsteady, three-dimensional intraglottal pressure loading in self-oscillating excised VFs, investigating the dependence of both anterior-posterior pressure gradients, and unsteady effects^57^. It was found that the intraglottal pressure reached negative gauge values only along the midline of the VFs, and not at the anterior and posterior edges. However, the diameter of the pressure sensors used to acquire the intraglottal pressure was relatively large (2.36 mm) compared to the inferior-superior and anterior-posterior VF dimensions (*O*(10 mm)), leading to poor spatial resolution of the flow and obfuscating some of the key dynamical behaviors.

Nevertheless, similar results have been observed using computational models of the VFs undergoing self-sustained oscillations^58,59^. Early efforts^3^ validated the computational model knematics with synthetic silicone VF models to investigate the aerodynamic energy exchange during oscillation. The results provided insight into the net energy transfer from the airflow to the VF tissue, which was found to be positive, validating a previously proposed theory that the energy transfer from the fluid to the VF must be greater during opening than closing to produce self-sustained oscillations^5^. However, modeling VF contact is highly challenging in numerical investigations, which can influence the accuracy of the oscillation dynamics.

Despite the extensive efforts devoted to resolving the intraglottal aerodynamic pressure during VF oscillation, temporal and spatial variations in the pressure have still not been accurately quantified. This deficiency arises from the challenging environment of voiced speech production, were tight geometric constraints, in tandem with the high-frequency of the VF oscillations, pose significant challenges such as accurate positioning of a probe between the VFs and the disruptive effect of the probe on the flow field^60^. Recently, a new approach that employs synthetic, self-oscillating VF models in a hemilaryngeal configuration was developed and validated for accurately measuring the intraglottal aerodynamic pressures during oscillation^60^. The self-oscillating VF model captures both three-dimensional and unsteady flow effects addressing the shortcomings of many prior works.

The objective of this work is to investigate the effects of flow unsteadiness and three-dimensionality on the intraglottal pressure distributions in a hemilaryngeal self-oscillating silicone VF model. The model is incorporated into a novel flow facility that enables, for the first time, both temporal and spatial resolution of the intraglottal pressure waveform during the opening and closing phases of the phonatory cycle. The pressure field is then used to investigate the energy exchange from the airflow to the VFs. The flow facility and the measurement procedure are described in Section II. The results and discussion are presented in Section III. Finally, Section IV is left for the conclusions.

## II. METHODS

### A. Hemilaryngeal Flow Facility

A synthetic, self-oscillating silicone model of the VFs was used in a hemilaryngeal flow facility to acquire the intraglottal pressure distribution in both the inferior-superior and the anterior-posterior directions. The geometry of the VF model and the experimental setup was similar to that used in prior work (See Section 3.1 in Motie-Shirazi et al.^60^), and is shown in Figure 1a.

**FIG. 1.**
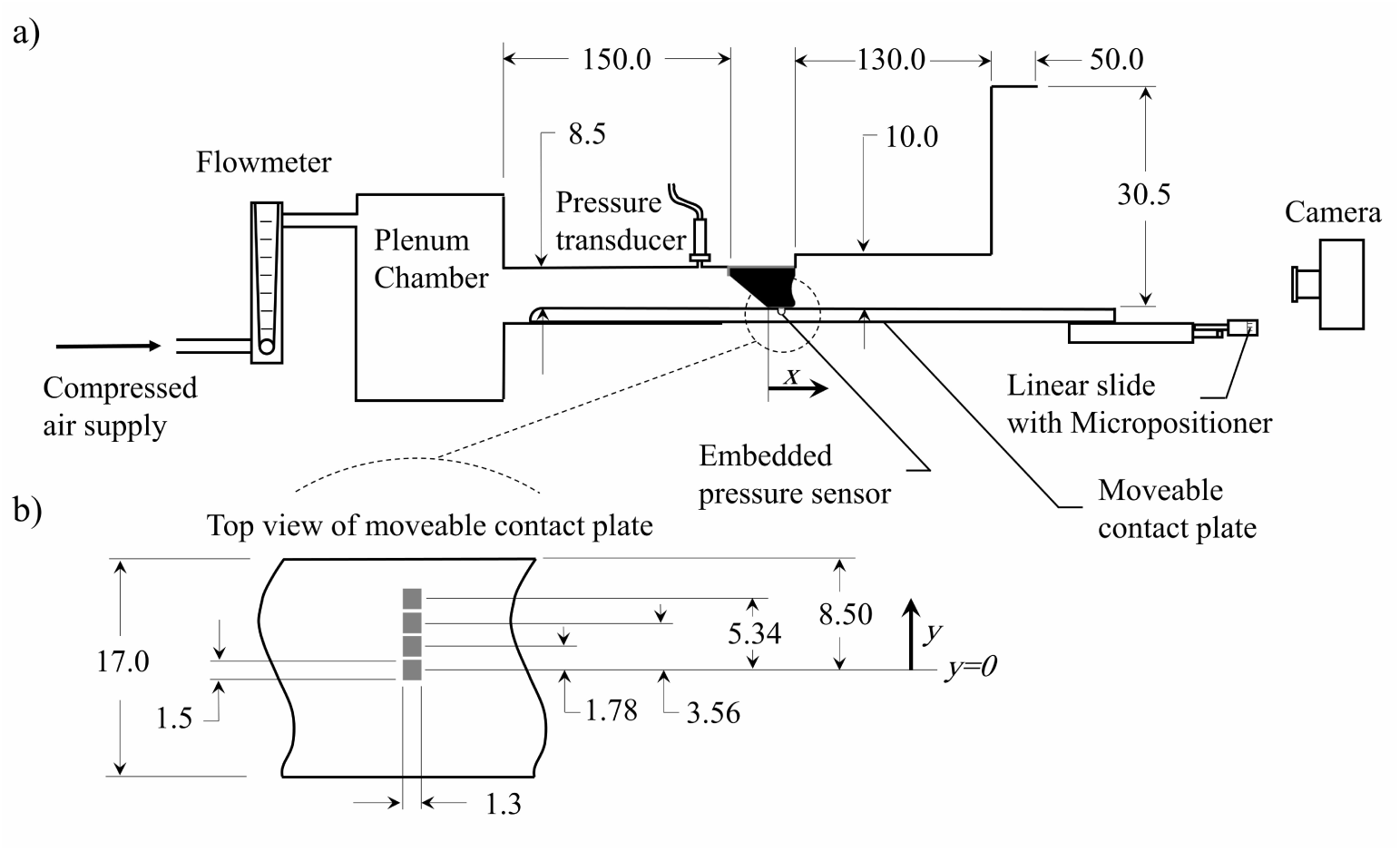
(a) Schematic of the experimental flow facility. (b) A close-up top view of the contact plate and the relative position of the pressure sensors. All dimensions are in mm.

Briefly, a constant pressure source provided flow through a Dwyer RMC 101-SSV inline flow meter (Dwyer, IN, USA) that measured the time-averaged flow rate. Upon exiting the flow meter, the flow entered a 0.03 m^3^ plenum chamber with a cross sectional area of 0.06 m^2^, whose inner walls were insulated acoustically by foam. The plenum chamber was connected to a model trachea that was comprised of a 213.0 mm^2^ area rectangular channel that was 150.0 mm long. The subglottal pressure was monitored with a Kulite ET-3DC pressure transducer (Kulite, NJ, USA). Together, the plenum chamber and square duct mimic the acoustic loading of the subglottal tract.

The VF model was mounted in a hemilaryngeal configuration by placing it in a bracket at the exit of the model trachea. The wall against which the VF oscillated extended the length of the tracheal tract, and could be moved in the inferior-superior direction. This is referred to as the contact plate (see Figure 1a). A Millar Mikro-Cath pressure sensor (Millar, TX, USA) was embedded in a groove in the contact plate. The groove was then filled with Smooth-On Dragon Skin 10 silicone (Smooth-On Inc, PA, USA) to provide a smooth level surface against which the VF model vibrated.

High-speed video (HSV) of the VF kinematics was acquired with a Photron AX200 charge-coupled device (CCD) camera (Photron, Tokyo, Japan) at 20, 000 frames-per-second and a resolution of 640 pixels by 480 pixels. The camera was positioned superiorly to the VF exit. An Elicar V-HQ Macro 90 mm f 2.5 lens (Jaca Corporation, Tokyo, Japan) provided a field of view measuring 48.0 mm by 36.0 mm. The HSV, subglottal pressure, and intraglottal pressure signals were all synchronized and acquired using a custom LabVIEW Virtual Instrument program (National Instruments Corporation, Austin, TX, USA). The subglottal and intraglottal pressure signals were recorded on a National Instruments PCIe-6321 data acquisition card (National Instruments Corporation, Austin, TX, USA) at 80 kHz for a duration of 0.75 s.

A vocal tract was added to the flow facility at the VF exit to include acoustic loading effects, as shown in Figure 1a. The geometry of the vocal tract was idealized based on the vocal tract geometry of the vowel /o/ reported from magnetic resonance imaging of the vocal tract^61^. The supraglottal channel had a constant width of 26.2 mm in the anteriorposterior direction, and a total length of 180.0 mm. The medial-lateral height consisted of two discrete, connected sections. The first section (exiting the glottis) had a length of 130.0 mm and a height of 10.0 mm (262.0 mm^2^ cross-sectional area), which then transitioned to the second section, which measured 50.0 mm long with a height of 30.5 mm (917.0 mm^2^ cross-sectional area).

Four identical contact plates were employed for the measurements, with each containing an embedded pressure sensor at a different location in the anterior-posterior direction; one at the anterior-posterior midline, with the subsequent ones equally spaced every 1.78 mm moving in the anterior direction (see Figure 1b). The coordinate system is defined such that *x* = 0 corresponds to the location of the inferior edge of the glottis when the VF is at rest and *x* values indicate the distance in the inferior-superior direction. In addition, *y* = 0 corresponds to the anterior-posterior midline and the *y* values indicate the distance in the anterior-posterior direction. A Thorlabs PT1 linear slide (Thorlabs, Newton, NJ, USA) moved the contact plate in the inferior-superior (*x*) direction to allow the pressure to be acquired at any location along this direction. The area over which the pressure was measured was 1.50 mm by 1.30 mm in the inferior-superior and medial-lateral directions, respectively. The linear slide had a positional accuracy of 0.0254 mm.

### B. Vocal Fold Model

The dimensions of the VF model, layer composition, and silicone mixture ratios used for each layer was the same as prior work^60^, with the dimensions and layer thicknesses shown in Figure 2. This profile was extruded in the anterior-posterior direction to a length of 17.0 mm.The modulus of elasticity of each silicone layer, however, was newly quantified using a TA Instruments AR 2000 Rheometer (TA Instruments, DE, USA), as opposed to the previously employed TA Instruments Q 800 dynamic mechanical analysis measurements (TA Instruments, DE, USA), to improve accuracy at the low moduli of interest. To perform the rheology measurements, cylindrical samples with a diameter of 60.0 mm and a thickness of 1.0 mm were created from the same batch of each silicone mixture as the VF layers. Elastic (*G*′) and viscous (*G*^″^) shear moduli were measured by performing a frequency sweep from 1 Hz to 100 Hz, which was the upper limit of the instrument, at 1% strain. The results are presented in Figure 3 and are compared with values from human VF cover measurements^62^. Good agreement with the physiological data is found for the silicone values.

**FIG. 2.**
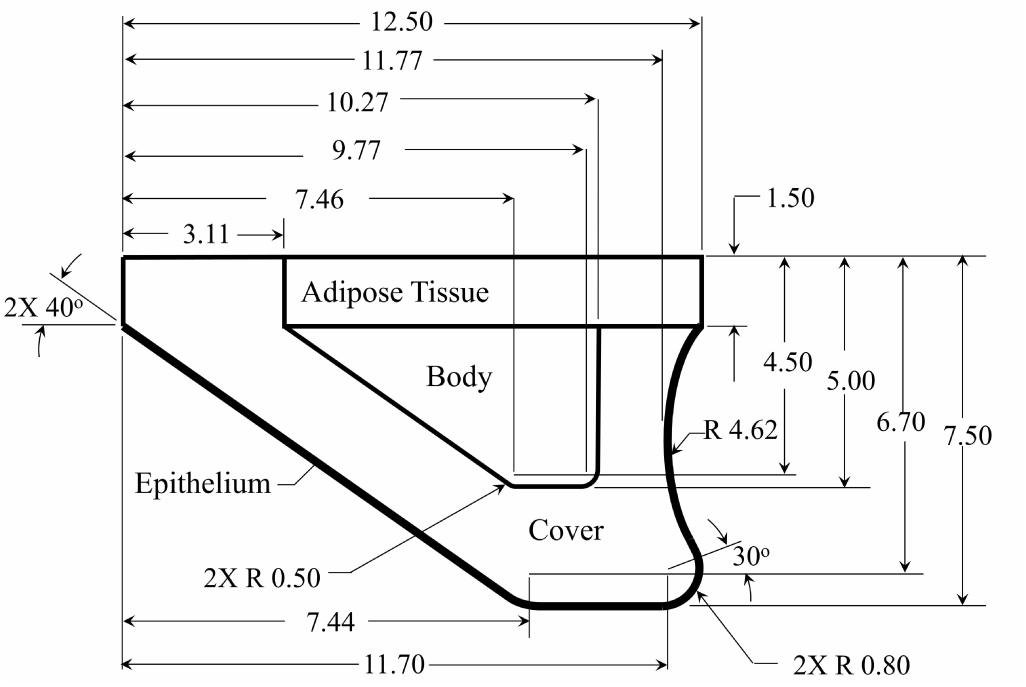
Geometry and dimensions of the synthetic VF model. All dimensions are in mm.

**FIG. 3.**
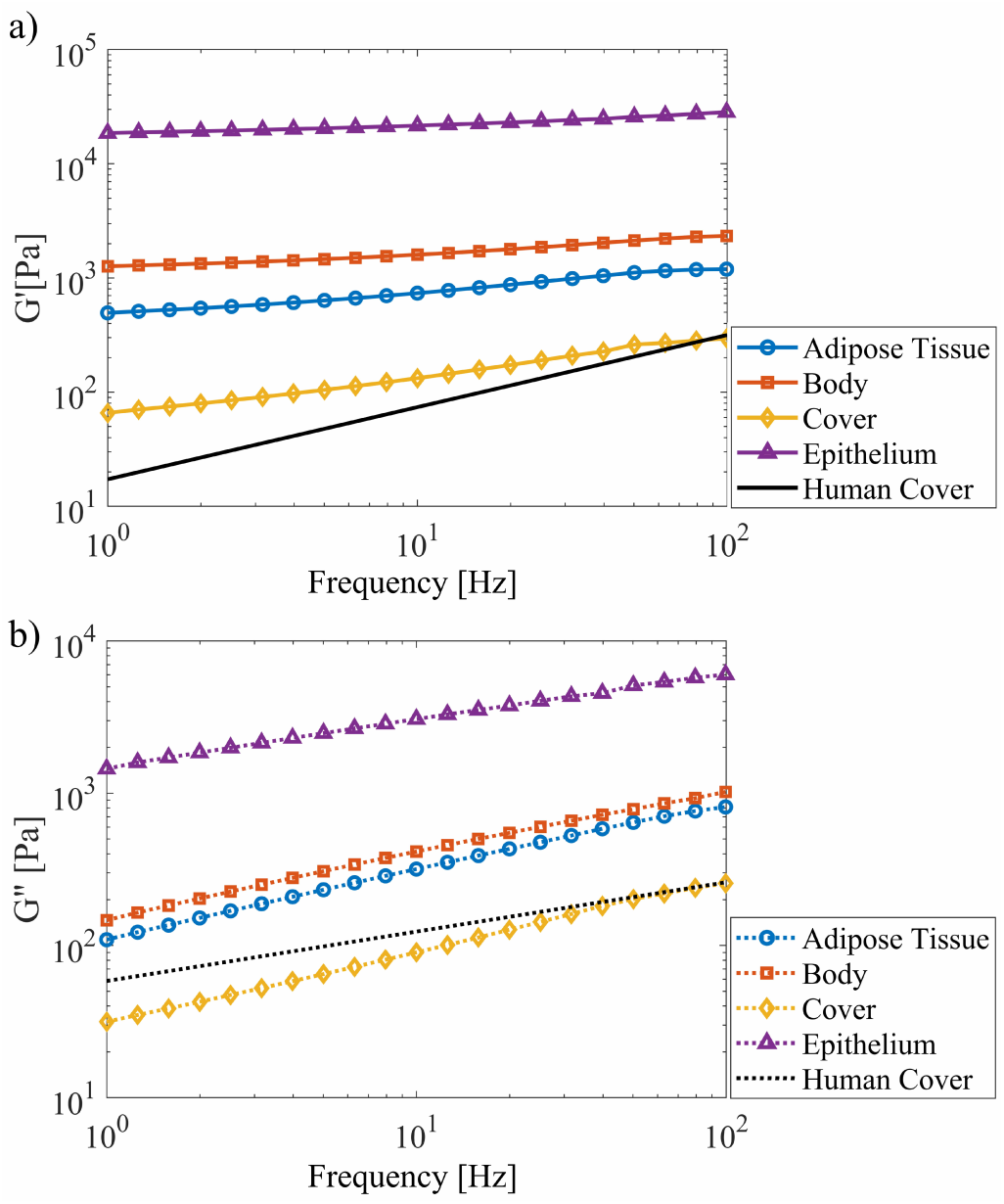
(Color online) (a) Elastic (*G*′) and (b) viscous (*G*^″^) shear moduli of different layers of the VF model with corresponding values for human VFs^62^. Axes are plotted on a logarithmic scale.

The complex modulus of elasticity (*E*^∗^) of an isotropic material is related to the complex shear modulus (*G*^∗^ = *G*′ + *iG*^″^) and Poisson’s ratio (*v*) by

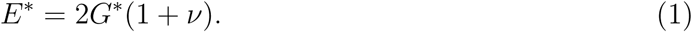

Poisson’s ratio has been measured to be ≈ 0.4 for different mixture ratios of silicone at 1% strain^63^. Using these relationships, the magnitude of complex modulus of elasticity at a frequency of 100 Hz was computed for each layer, and is reported in Table I alongside the range of physiological values. Good agreement is found between the physiological and synthetic values.

**TABLE I.** Moduli of elasticity of physiological and silicone vocal fold models for each layer.

| Layer | Physiological range<br>(kPa) | Silicone VF Model<br>(kPa) |
| --- | --- | --- |
| Adipose tissue | $1 - 10^{79}$ | 4.04 |
| Body | $1.5 - 50^{80-82}$ | 7.13 |
| Cover | $1 - 8^{62,81,83-86}$ | 1.10 |
| Epithelium | Not measured | 81.10 |

### C. Pressure Measurement Procedure

During VF oscillation each of the movable contact plates was independently inserted and positioned such that the pressure sensor was located superior to the VF. The contact plate was then moved in the inferior direction in increments of 0.254 mm. The unsteady intraglottal pressure at each location was acquired by the pressure sensor. Figure 4 shows the initial and final positions (I and II) of the sensor relative to the VF model at rest. Note that the airflow caused the VF to bulge slightly in the superior direction such that contact did not occur at the exact glottal midpoint of the rest position of the VF model. After sampling for 0.75 s at each location, which produced about 120 cycles for an oscillation frequency of 160 Hz, the waveforms were then phase-averaged based on the oscillation cycle, resulting in a time-varying mean pressure waveform.

**FIG. 4.**
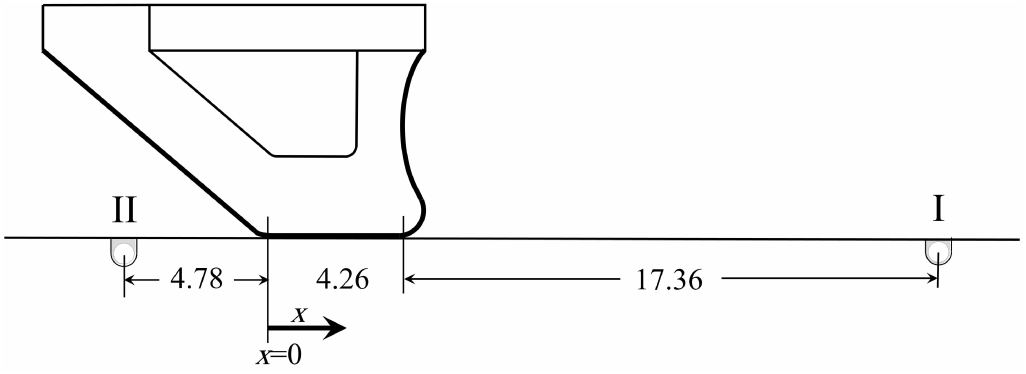
Schematic of the initial (I) and final (II) positions of the pressure sensor moving with the contact plate relative to a stationary VF model. All dimensions are in mm.

## III. RESULTS AND DISCUSSION

### A. Oscillation Dynamics

VF oscillation dynamics were investigated with a medial prephonatory compression of 0.75 mm that resulted in a medial prephonatory pressure of 1.25 kPa. These are, respectively, the medial distance that the VF is deformed into the hemilaryngeal plate when it is in a static configuration, and the resulting static pressure (see Motie-Shirazi et al.^60^ for further details). Due to the medial compression, the length of the glottis in the inferior-superior direction during self-oscillation was ∼ 1.5 times greater than for the static geometry with no medial compression. The onset pressure of the VF model was measured to be 1.70 kPa. The intraglottal pressure measurements were performed at a mean subglottal pressure of *p*_sub_, = 2.20 kPa. The mean flow rate was measured to be 338 mL*/*s. The fundamental frequency of oscillation was 160 Hz yielding a period (*T*) of 6.25 ms.

Figure 5 shows a kymogram plot of an oscillation cycle at the mid anterior-posterior location extracted from the HSV. At this location, the open quotient (*OQ* = *T*_open_,*/T*) was calculated to be ≃ 0.78 and the speed quotient (*SQ* = *T* ^+^ */T*^−^) was ≃ 2.09 where *T*_open_,, *T, T* ^+^, and *T*^−^ are defined in Figure 5. Both *OQ* and *SQ* are in the physiological ranges for human VFs^64,65^. The visible medial boundary of the VF was determined by defining a threshold value, and a least-squares regression^66^ was then used to fit two sinusoidal functions to the boundary during the VF opening and closing phases, indicated by the solid and dashed lines, respectively. These functions were then used to estimate the VF surface velocity to calculate the aerodynamic energy, as explained in Section III B 3. The onset pressure and flow rate of the synthetic model were higher than physiological values^67^. This behavior has been observed in prior investigations with hemilaryngeal configurations^57,68,69^. In addition, a relatively high medial prephonatory compression was required to get robust contact in the synthetic models, which also increased the onset pressure and the flow rate. In spite of the higher subglottal pressures, the VF kinematics are representative of physiological motion.

The glottal height (*d*_g_,) at the four positions in the anterior-posterior direction are plotted in Figure 6 as a function of time (*t*), normalized by *T*. The distance from the midline, *y* (See Figure 1b), is normalized by the glottal half width in the anterior-posterior direction, which was *w*_g_,*/*2 = 0.85 mm. The physical locations relative to the model geometry are indicated in the inset image. Note, the stepwise behavior of the glottal distance is due to the limit of the spatial resolution of the images, which was 0.077 mm*/*pix. The maximum glottal distance occurred at the midpoint and was calculated to be *d*_g,max_, = 0.67 mm, which is physiologically accurate^70^. It can be seen that VF closure occurred at the sides first, and then progressed toward the midline, with a similar pattern observed during opening. Moreover, the open quotient was greatest at the midpoint, and decreased towards the anterior-posterior endpoints. The maximum glottal area was 8.45 mm^2^, which is also representative of clinical measures^71^.

**FIG. 5.**
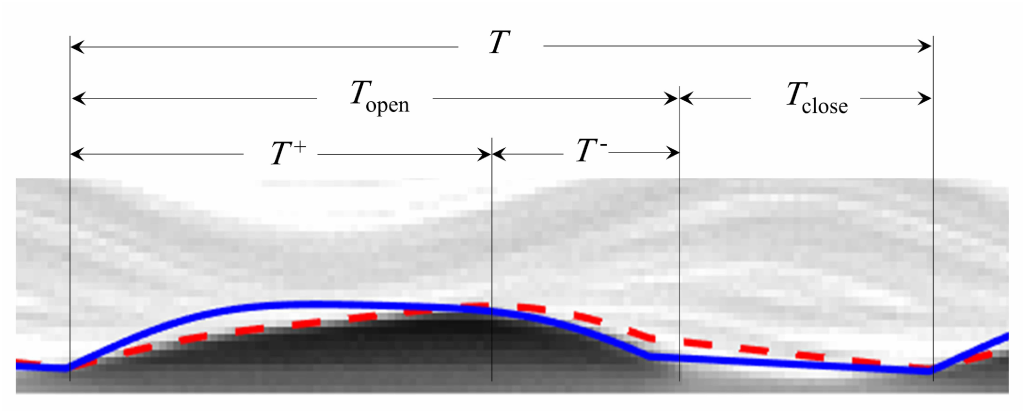
(Color online) Kymogram at the mid anterior-posterior location of the VF model. Solid and dashed lines identify the sinusoidal functions that were respectively fitted to the inferior edge of the VF during closure, and the superior edge during opening, via least-squares regression^66^.

**FIG. 6.**
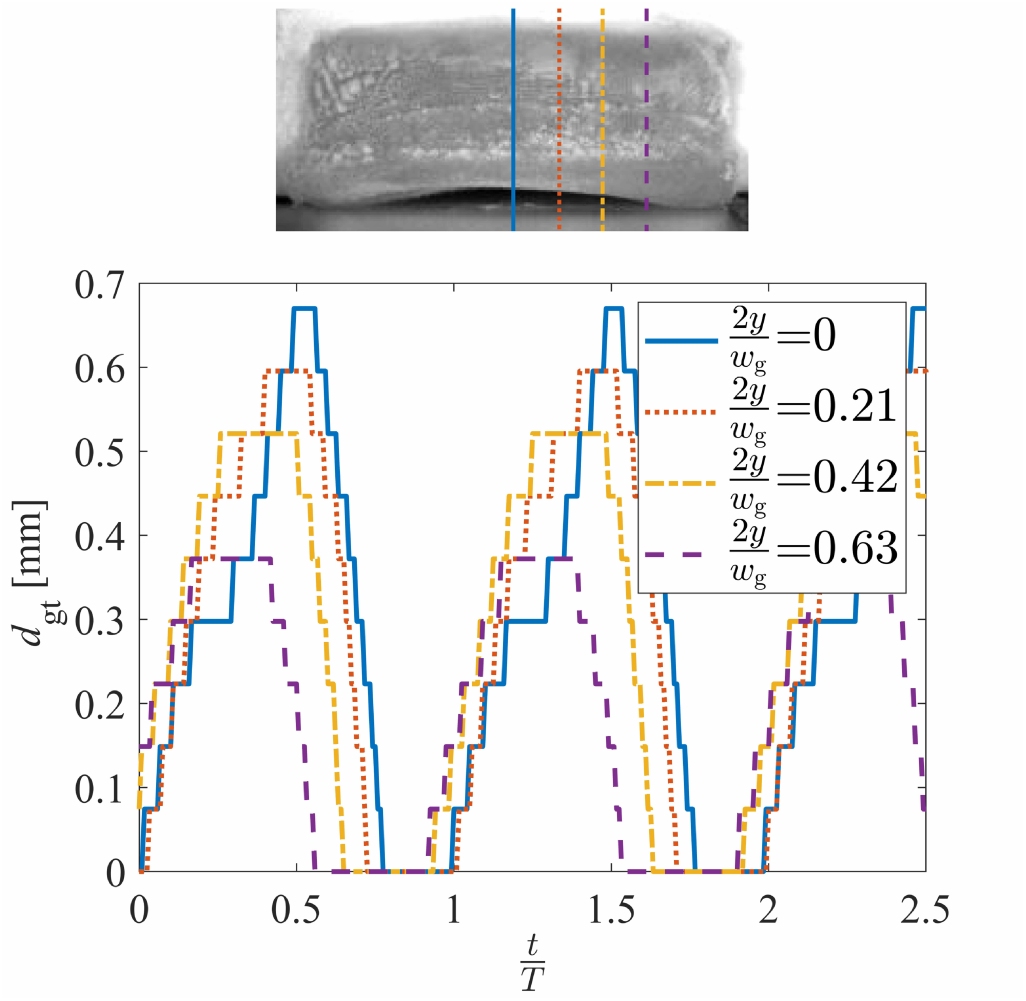
(Color online) Glottal distance at different anterior-posterior locations as a function of normalized time.

### B. Aerodynamic Pressure

#### 1. Convergent Phase

The spatial variation of the intraglottal aerodynamic pressure in the inferior-superior direction was computed as the normalized pressure drop, relative to the subglottal pressure, *p*_sub_,, expressed as (*p* − *p*_sub_,)*/p*_sub_,. Figure 7(a-d) presents surface pressure measurements at the four anterior-posterior locations and at four different instances in time during VF opening, corresponding to *t/T* = 0, 0.26, 0.39, and 0.45, respectively. The abscissa indicates the distance from the inferior glottal margin when the VF is at rest and the medial prephonatory compression is not applied, normalized by the inferior-superior glottal length of *l*_g_, = 4.26 mm (See Figure 4). The inset images are a superior view of the VF orientation at the corresponding times, and denote the anterior-posterior position of the four pressure measurements, indicated by the corresponding line types. At the VF midline, the opening started at *t/T* = 0 and ended at *t/T* = 0.52. The dashed vertical lines *x/l*_g_, = 0 and *x/l*_g_, = 1 indicate the location of the inferior and superior boundaries of the glottis when the VF is at rest.

**FIG. 7.**
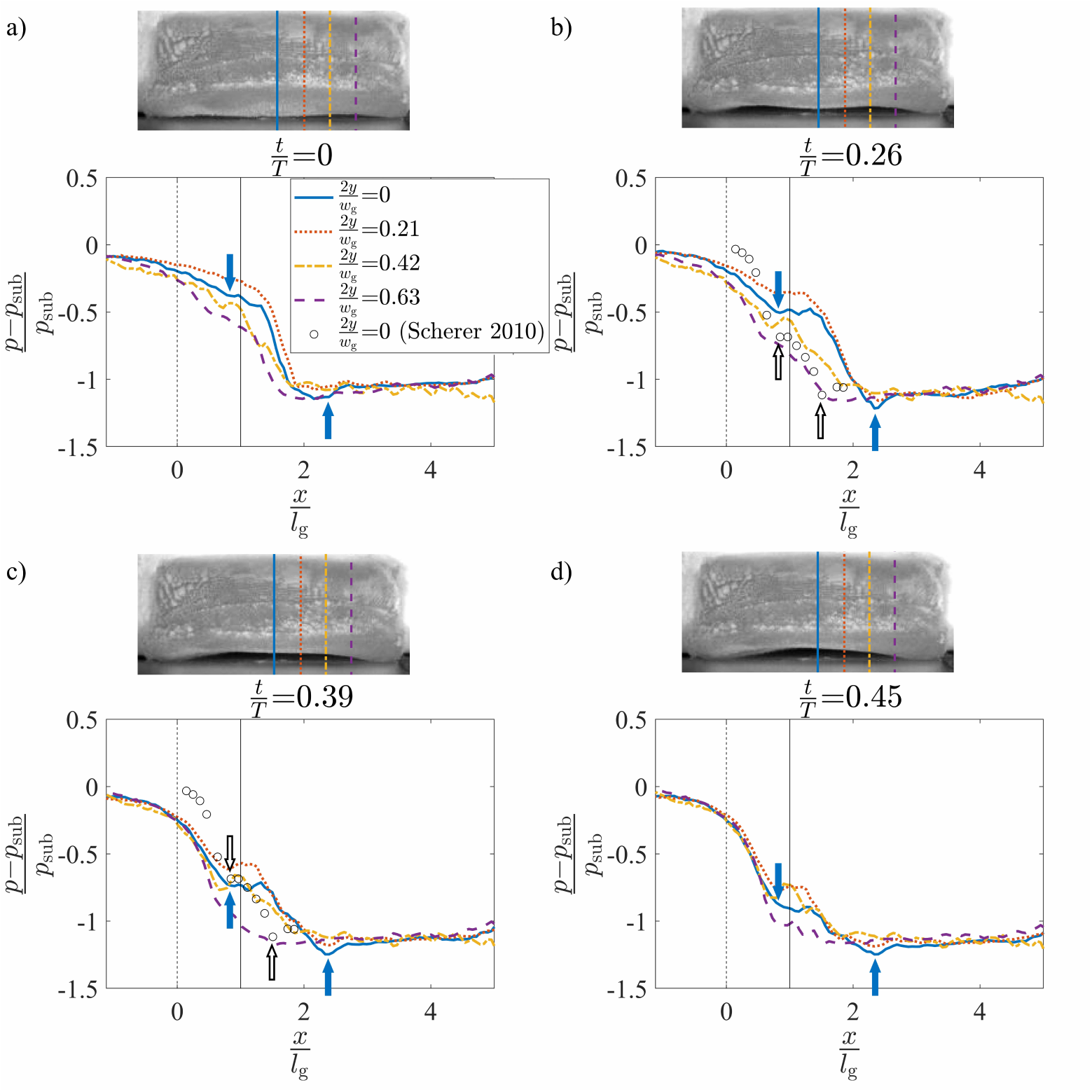
(Color online) The normalized intraglottal pressure drop versus the normalized inferior-superior distance, plotted at four positions in the anterior-posterior direction, and at normalized times of (a) *t/T* = 0, (b) *t/T* = 0.26, (c) *t/T* = 0.39, and (d) *t/T* = 0.45. The times coincide with a convergent glottal configuration. The solid arrows indicate the glottal entrance and exit at 2*y/w*_g_, = .0. Each inset presents a superior view of the VF orientation at the same instance in time. Dashed and solid vertical lines at x=0 and x=1, respectively, identify the glottal entrance and exit when the 0.0. Each inset presents a superior view of the VF orientation at the same instance in time. Dashed and solid vertical lines at x=0 and x=1, respectively, identify the glottal entrance and exit when the VF is in its rest configuration. Pressure measurements from steady flow, static investigations^56^ are included in subplots (b) and (c) with the glottal margins indicated by hollow arrows.

A video showing the progression of the intraglottal pressure distributions throughout the entire oscillatory cycle is included as a supplementary file^72^. The still images of Figure 7 are extracted from this video, as well as those discussed in Section III B 2 for the divergent orientations.

During VF opening (see Figure 7) the mean subglottal pressure decreased as the flow accelerated as it entered the glottis. A sharp decrease in the pressure occurred as the flow passed through the glottis. Note that for Figure 7(b-d) the pressure within the glottal region plateaus, before sharply decreasing. This can be explained by the viscous losses within the glottis being balanced by the total acceleration of the flow. The minimum pressure then occurred at the glottal exit, where the glottal gap height was smallest. The location of the glottal entrance and exit along the midline of the VF are noted in Figure 7 by the successive solid arrows. Comparing these locations across Figure 7(b-d) shows that the glottal entrance and exit were largely stationary during the opening phase of the VF oscillation. Nevertheless, the consistent shift in the location of the superior edge of the VF (the minimum pressure location) relative to the static position of the superior VF edge indicates the presence of superior VF bulging during oscillation. The intraglottal pressure measurements were compared with prior results acquired along the midline of three-dimensional static VF models in steady flow^56^, represented as circles in Figure 7(b,c). The static models had a 10^°^ convergence angle and a fixed medial-lateral glottal distance of 0.8 mm. For comparison, the estimated location of the glottal entrance in the current self-oscillating model was aligned with the glottal entrance in the prior static model investigations. The inferior-superior distance of both the prior static model investigations and the current self-oscillating ones were normalized by each corresponding inferior-superior glottal length, which were 3.00 and 4.26 mm, respectively. The glottal entrance and exit of the static model are indicated with hollow arrows in Figure 7(b-c). The location of the glottal exit did not match because the glottal length of the synthetic model increased during oscillation, as described above, but was fixed for the static model. The intraglottal pressure profiles in these models are very similar, with a change in the slope at the entrance and a drop in the pressure at the glottal exit. Similar behavior has been reported in static models with convergent glottal profiles^31,35,36,40,73^.

The effect of unsteady flow was investigated by comparing the temporal progression of the intraglottal pressure waveform during VF opening, presented sequentially in Figure 7(a-d). The mean intraglottal pressure decreased at all anterior-posterior locations as the VF was opening. This contradicts the oft-employed quasi-steady assumption that is invoked when investigating pressure-flow relationships through static VF models. As the glottal area increases during opening, the quasi-steady assumption predicts the glottal pressure should increase. Instead, the increased subglottal pressure that builds up during VF closure results in a temporally-accelerating flow field that produces a significant decrease in the nondimensionalized intraglottal pressure drop. While prior work has identified the existence of high flow acceleration during opening^17^, these findings indicate how the accelerating flow influences the intraglottal pressure field. In particular, these results show that the decrease in pressure due to flow acceleration is much greater than the pressure increase associated with the increasing glottal area.

The average intraglottal pressure varied in the anterior-posterior direction as a function of time. During the later stages of opening (Figure 7(c-d)), the pressure magnitude in the entrance of the glottis remained largely constant, while the more anterior positions of 2*y/w*_g_, = 0.21 and 0.42 increased slightly, before subsequently decreasing. At 2*y/w*_g_, = 0.63 the pressure is significantly lower for all inferior-superior positions. These variations are likely due to the three-dimensional geometry, which leads to varying pressure losses within the glottis. This, in turn, drives three-dimensional flow behavior, as has been previously observed^56^. Moreover, as can be seen in Figure 7, the pressure field at the anterior-posterior location of 2*y/w*_g_, = 0.42 exhibited significantly different behavior than that observed at the midline. During the early phases of opening, the pressure was lower than that at the midline, but then increased to higher values as the cycle progressed. This is likely due to the anterior-posterior asymmetry in VF closure, where the midline closes slightly sooner than more anterior locations.

#### 2. Divergent Phase

The spatial variation of the normalized intraglottal pressure drop during VF closure is plotted in Figure 8(a-d) at four normalized times of *t/T* = 0.52, 0.68, 0.74, and 0.77, respectively. The pressure profiles are not presented at 2*y/w*_g_, = 0 and 0.21 because closure at these locations has already occurred at the selected times. Similarly, 2*y/w*_g_, = 0.21 is not plotted in Figure 8(d) for the same reason. The inset images show a superior view of the VF motion at the same instant in time, with the vertical lines indicating the anterior locations at which the aerodynamic pressure was recorded. A clear divergent profile was present during the closing phases of oscillation, as was observed in the kymogram of Figure 5) and a kymogram extracted from the position 2*y/w*_g_, = 0.21 (not shown for brevity). For kymograms extracted at the more anterior/posterior positions of 2*y/w*_g_, = 0.42 and 0.63, propagation of the mucosal and the resultant divergent profile were not as pronounced.

**FIG. 8.**
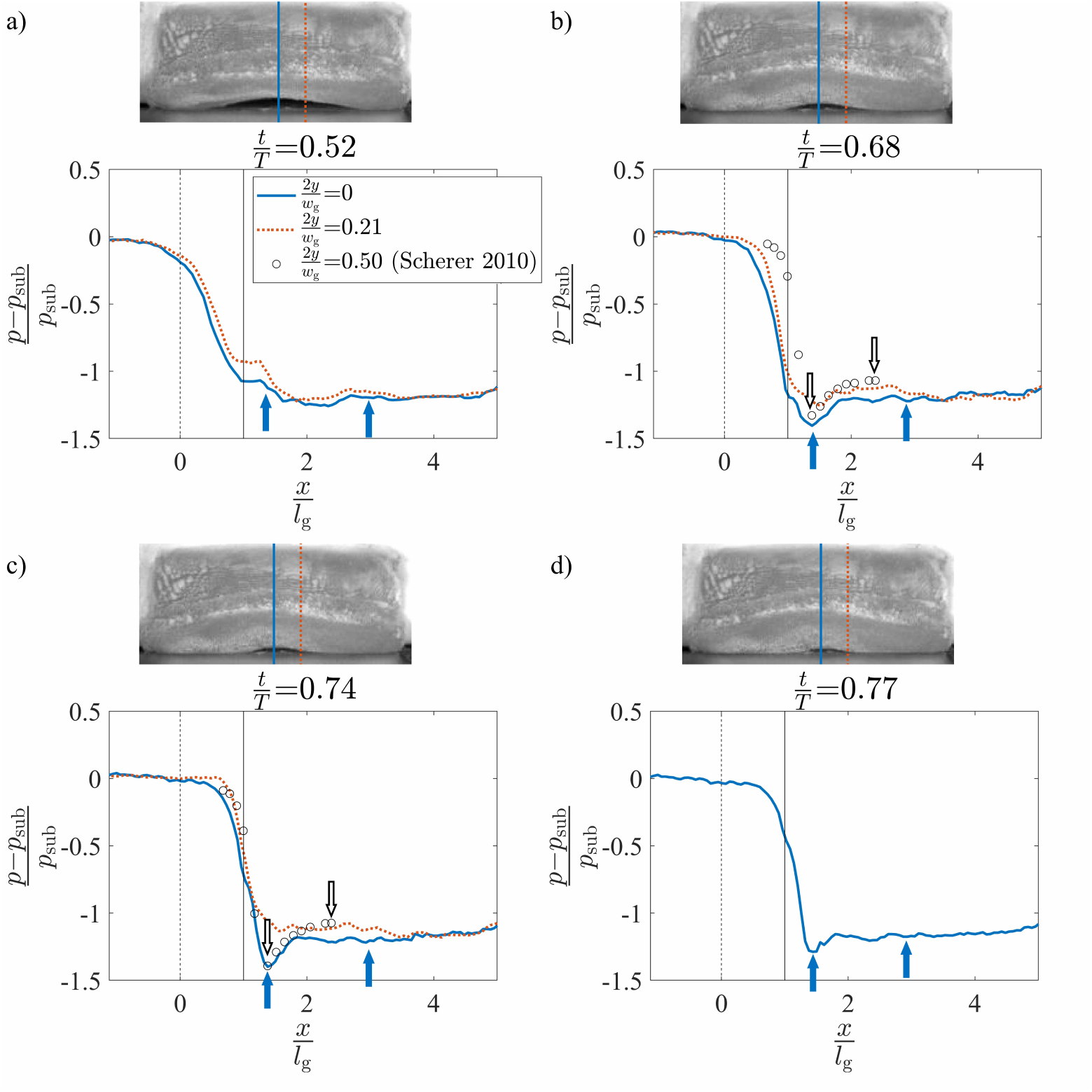
(Color online) The normalized intraglottal pressure drop versus the normalized inferior-superior distance, plotted at four positions in the anterior-posterior direction, and at normalized times of (a) *t/T* = 0, (b) *t/T* = 0.26, (c) *t/T* = 0.39, and (d) *t/T* = 0.45. The times coincide with a divergent glottal configuration. The solid arrows indicate the glottal entrance and exit at 2*y/w*_g_, = 0.0. Each inset image presents a superior view of the VF orientation at the same instance in time. Dashed and solid vertical lines at x=0 and x=1, respectively, identify the glottal entrance and exit when the VF is in its rest configuration. Pressure measurements from steady flow, static investigations^56^ are included in subplots (b) and (c) with the glottal margins indicated by hollow arrows.

The glottal entrance and exit at the midline of the VF are indicated by the solid arrows in Figure 8(b-d). The entrance locations were identified based on the assumption that the pressure will be a minimum at the glottal entrance (the minimum glottal area) when the glottis assumes a divergent orientation. At the beginning of VF closure when *t/T* = 0.52 (Figure 8(a)), the glottis was largely a uniform channel, and so it was difficult to identify the precise location of the glottal entrance. In this state, the glottal entrance was presumed to be at the same position as found at the other times during closure because, as can be seen in Figure 8(b-d), this location did not change significantly over time. Downstream of the minimal glottal area, pressure recovery occurred in the divergent glottal channel. The precise location of the glottal exit was difficult to identify because at the exit of the glottis, the pressure was essentially the same as the pressure in the supraglottal tract. Therefore, the location of the glottal exit at each of the anterior locations was estimated by assuming that the glottal length remained equal to the value found for the convergent glottal orientation, as described in Section III B 1.

From the pressure distributions it can be seen that the glottal entrance at the anterior-posterior midline of the VF was shifted to *x/l*_g_, ≃ 1.4. This indicates increased superior bulging relative to the convergent phase, where the glottal entrance was at *x/l*_g_, ≃ 0.8. The minimum pressure in the divergent phase was lower than in the convergent phase due to the pressure recovery arising from the divergent orientation.

The intraglottal pressures are compared in Figure 8 with prior results of steady flow through three-dimensional static VF models with a 10^°^ divergent glottal profile and a fixed medial-lateral glottal distance of 0.8 mm.^56^ The intraglottal pressure distributions obtained from the static model investigations at anterior positions of 2*y/w*_g_, = 0.50 and 0.0^56^ are plotted as hollow circles in Figure 8(b, c), respectively. For comparison, the divergent angle of the self-oscillating VF model was estimated from the recorded HSV during the closing phase of the VF based on the known medial-lateral displacement of the inferior and superior edges (see the VF edges at *T*^−^ in Figure 5), and the inferior-superior length of the medial surface when at rest. It was found that at an anterior position of 2*y/w*_g_, = 0.21, a divergent angle of ∼ 10^°^ occurred at *t/T* = 0.68 (Figure 8(b)) and for an anterior position of 2*y/w*_g_, = 0 the divergence angle was ∼ 10^°^ at *t/T* = 0.74 (Figure 8(c)). Similar to Figure 7, the estimated location of the glottal entrance was aligned with the glottal entrance from the static model investigations, and the inferior-superior length of each scenario was normalized by the inferior-superior medial surface length. Reasonable agreement is observed between the static and dynamic pressure profiles, which is not surprising as unsteady effects due to flow acceleration are not anticipated to be as significant during the latter phases of the phonatory cycle. The magnitude of the minimum pressure was lower at the middle when compared to the anterior/posterior positions, again indicating the presence of three-dimensional flow behavior.

The temporal variation of the intraglottal pressure during VF closure was observed by comparing the temporal evolution of the minimum pressure peak, which occurs at the minimal glottal area. As the VFs closed, this value decreased as the minimum glottal area became smaller, and the subsequent pressure loss across the glottis increased. However, it is interesting to note that immediately preceding closure, at *t/T* = 0.77, the minimum pressure increased, despite the continued decrease in the glottal area (see the insets of Figure 8(c,d)). This increase in the minimal pressure can be explained by an increase in the viscous pressure loss when the glottal orifice becomes very small at the end of closure, resulting in a decrease in the flow velocity and consequently a rise in the pressure. This is an important observation that is not captured by static VF investigations.

#### 3. Aerodynamic Energy

The transfer of energy from the airflow to the VF within the glottis was investigated. The power transferred to the VF is approximately equal to the product of the normal force applied on the VF glottal surface and the normal component of the surface velocity^3^. The VF velocity magnitude in the inferior-superior direction is negligible compared to the medial-lateral velocity and the viscous forces applied on the VF have a minor influence on the transferred power^3^. Therefore, the medial-lateral component of the velocity and the fluid pressure were used to estimate the power transfer. At each of the four anterior-posterior locations, the medial-lateral component of glottal surface velocity was estimated from kymogram plots at the same position where the pressure measurements were acquired. Two sinusoidal functions were fit to the visible medial boundary of the VF, which is the superior edge during opening and the inferior edge during closing, using least-squares regression^66^, as shown in Figure 5. The magnitude of the glottal surface velocity in the medial direction (*v*_g_,) was then estimated by computing the derivative of these functions at each instance in time. This assumes the medial VF surface velocity is equal to the superior edge velocity during opening, and the inferior edge velocity during closing. While admittedly an approximation, it provides a first-order estimate of the VF surface velocity. Because the VF surface velocity could not be estimated outside the glottal region, the power transfer was only investigated within the glottis.

The medial VF displacement and surface velocity are presented in Figure 9(a) as a function of the normalized time over an oscillation cycle. The lines with circle symbols represent the medial-lateral glottal distance during opening and closure, with the ordinate axis on the left showing the magnitude. The lines without symbols are the corresponding medial surface velocity, as represented by the ordinate axis on the right. The dash-dot line identifies the contact phase when the VF was closed and the displacement and the velocity were zero. The velocity of the medial surface was positive during opening and negative during closure.

**FIG. 9.**
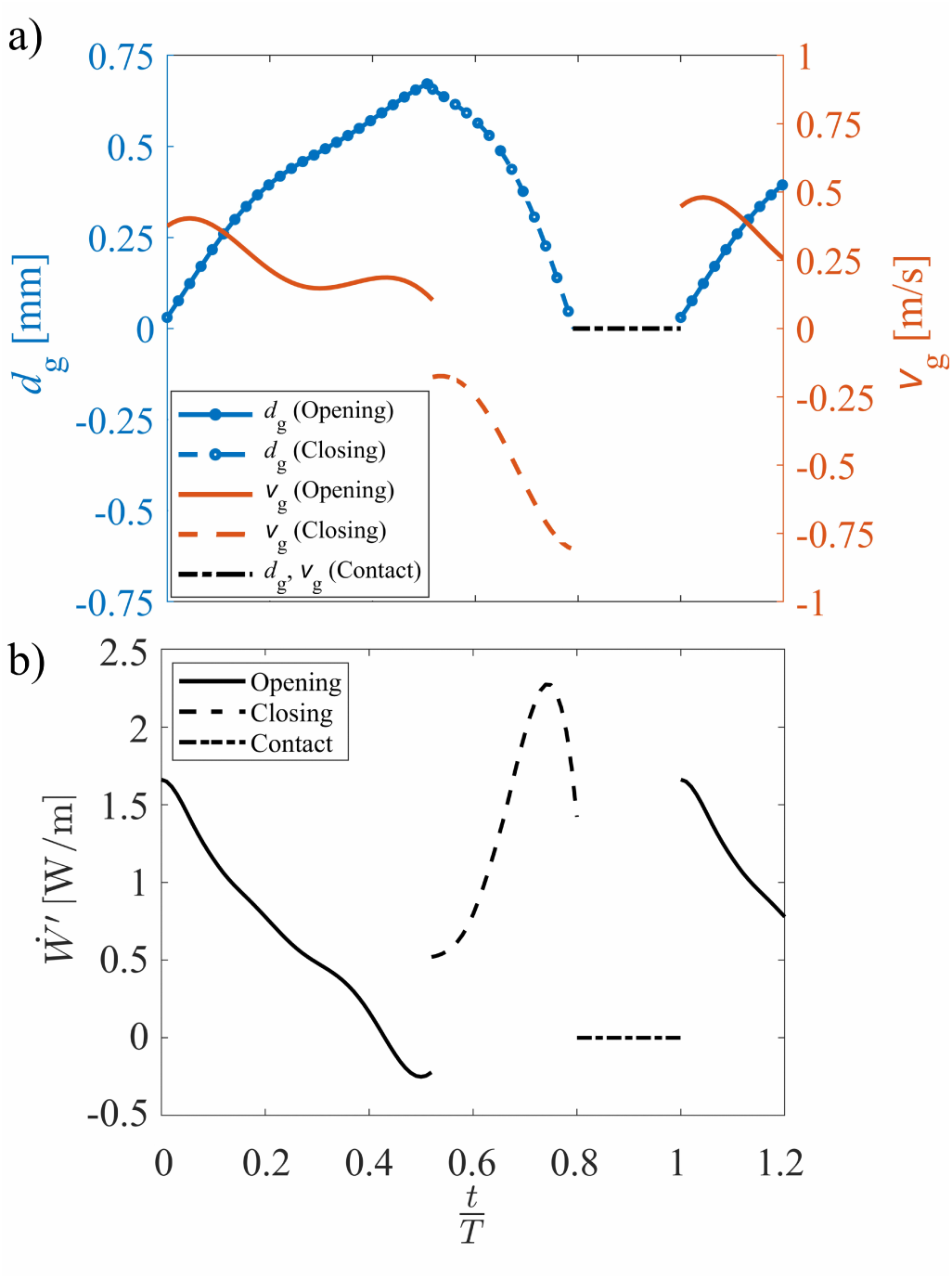
(Color online) Temporal change in (a) the glottal distance and velocity calculated from the fitted curves to the extracted kymogram, and (b) the power per unit anterior-posterior width, at 2*y/w*_g_, = 0.0 during the VF opening phase, closing phase, and contact.

At each of the anterior-posterior locations, the time-varying aerodynamic power per unit anterior-posterior width 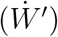, denoted by the prime superscript, during opening and closing of the VF was calculated by multiplying the VF surface velocity with the integral of the pressure between the inferior andsuperior margins of the glottis according to

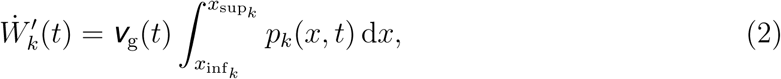

where *p*(*x, t*) indicates the intraglottal pressure at the inferior-superior location of *x* at time *t*. The limits *x*_inf_, and *x*_sup_, denote the location of the inferior and superior edges of the VF, which were estimated based on the pressure distributions during the opening and closing phases, using the method explained in Sections III B 1 and III B 2. The subscript *k* varies from 1 − 4 and corresponds to the four anterior-posterior positions at which the pressure was acquired.

The time dependence of 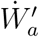 at the midline of the VF is plotted in Figure 9(b). During VF opening the intraglottal power was initially positive because the spatial average of the intraglottal pressure was in phase with the surface velocity and they both have a positive magnitude, indicating the transfer of energy from the airflow to the VF. The power decreased in time due to the decreasing intraglottal pressure, as can be seen in the sequential progression of Figure 7(a-d). This was due to the unsteady flow acceleration. The power transfer became negative during the last 10% of the opening phase as the spatial average of the intraglottal gauge pressure became negative while the surface velocity was still positive, denoting that energy began to be transferred from the VF back to the fluid even as the VF was still opening. This is a unique behavior that arises from the unsteady flow acceleration, and that is not captured by quasi-steady investigations.

A discontinuity is observed in Figure 9 when the VF transitioned from the opening to closing phase. This is due to the discrete change in the surface velocity that occurred due to the technique that was employed for estimating it. Namely, the assumption that the medial surface velocity is equal to velocity at the superior VF edge during opening, and the inferior edge during closing.

During the closing phase, both the intraglottal pressure and velocity were negative, which lead to a positive power transfer, indicating that the direction of the energy transfer was again from the fluid to the VF. The power initially increased during the closing phases as the intraglottal pressure became more negative as the glottal orifice became smaller and the flow accelerated (see Figure 8(b, c)). Immediately preceding closure, however, the power rapidly decreased due to the rapid increase in the intraglottal pressure, as was observed in Figure 8(d), likely because of the decrease in the glottal velocity due to the high viscous losses when the glottal orifice becomes very small, immediately preceding contact.

Although the mechanics of energy transfer have been studied numerically^5,53,74^, computationally^3^, and in driven synthetic VF models^16^, they have never been explored experimentally in self-oscillating VFs. Nevertheless, the calculated aerodynamic power variation can be compared with values from the prior investigations utilizing two-dimensional computational models^3^ and a driven synthetic VF model^16^. In both of these computational and driven model studies, the power was computed over the entire medial surface of the VF, as opposed to considering only the contribution from the medial component of the VF surface velocity and only in the glottal region, as performed in the current study. Nevertheless, similar behavior was observed in the time-varying aerodynamic power during the opening phase of oscillation, with an initial positive value that subsequently decreased, reaching a negative value as the glottis reached maximum opening (compare Figure 9(b) to Figure 12 in^3^ and Figure 13 in^16^). However, during the closing phase, the power in the computational and the driven model investigations reached more negative values at the beginning of the closing phase and then increased, becoming positive only at the end of the closing phase. In the current study the power was always positive during the closing phase. This discrepancy is likely due to the power transfer that occurred in the VF entrance regions, which was included in the computational and the driven models and not in the current synthetic silicone model investigation. For example, at the beginning of the closing phase, the magnitude of the pressure within the glottis was negative while the pressure in the subglottal region was positive, as can be seen in Figure 8(a). This resulted in positive power transfer in the self-oscillating model, which only considered the negative pressure within the glottis when computing the power transfer. In contrast, the power transfer in the computational and the driven models were positive because it considered the pressure distribution across the entire VF surface, including the regions of positive pressure inferior to the glottis. However, as the VFs closed, the intraglottal pressure decreased, see Figures 8(c,d). Because the surface velocity inside the glottis is higher than along the inferior surfaces, the power transfer that occurs within the glottis drives the energy exchange. This is evidenced by the net power subsequently increasing in all of the models. One important feature of the power transfer that was not captured by the computational and the driven models was the sharp drop in power immediately preceding VF closure, which was due to increased pressure losses arising from viscous effects as the glottal orifice becomes very small. This is likely because in the computational investigations, the collision phase was not modeled (i.e., a finite glottal area was always present) thereby altering the aerodynamic behavior during closure. In the driven model, a Bernoulli flow assumption was used to obtain the pressure field, which neglected the viscous effects of the flow and was not able to predict the pressure rise at the end of the closing phase.

The variation of the intraglottal aerodynamic energy transfer as a function of anterior-posterior position was also investigated. The aerodynamic energy transferred per unit width (*W*′) during the opening and closing of one VF oscillation was calculated by integrating the power per unit width over time as

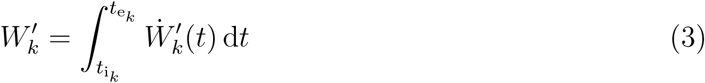

where *t*_i_, and *t*_e_, are the times corresponding to start and end of the opening or closing of the VF at the specified anterior-posterior location, as denoted by the subscript *k*. The total aerodynamic energy per unit width at the four anterior-posterior directions is shown in Figure 10 along with the respective contributions from the opening and closing phases. The total aerodynamic energy transfer was positive across the entire anterior-posterior width, indicating that throughout an oscillation cycle, net energy is transferred from the fluid to the VF across the anterior-posterior direction. However, the total energy per unit width decreased significantly from the midline to the anterior edge, indicating that the majority of the energy exchange occurs at the middle of the VFs. The total energy at the location of 2*y/w*_g_, = 0.63 was only 14% of the value at the midline. These findings further reinforce the importance of considering three-dimensional effects.

**FIG. 10.**
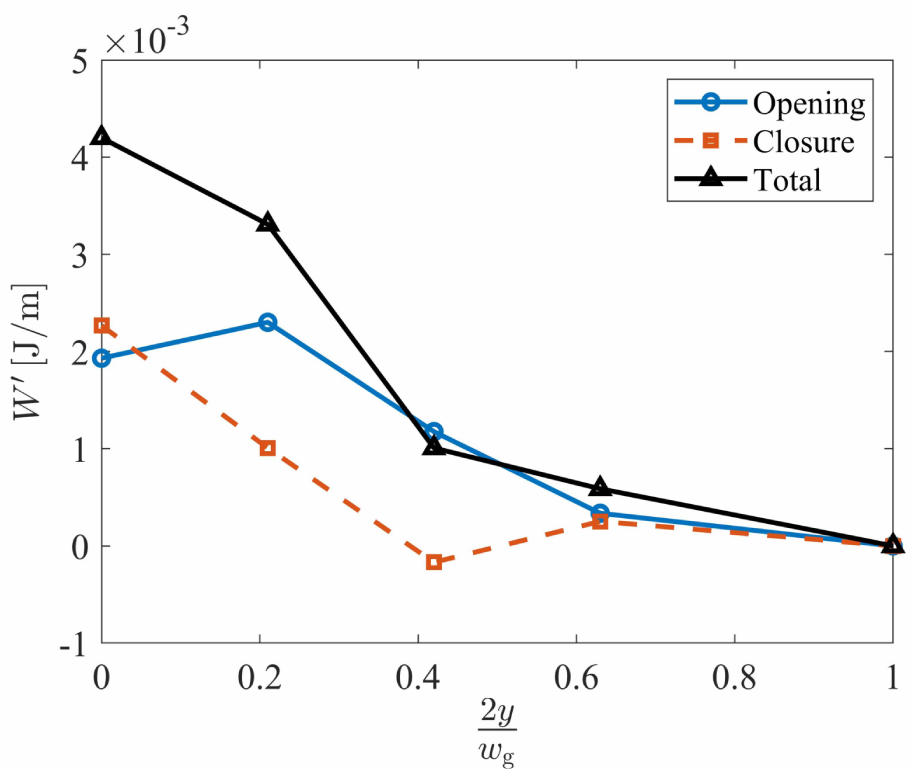
(Color online) Aerodynamic energy per unit anterior-posterior width computed during VF opening and closing, and the total aerodynamic energy transferred over the entire phonatory cycle, as a function of anterior-posterior location.

It is interesting to note that while the midpoint experienced largely the same amount of positive energy transfer during both opening and closing, the more anterior locations exhibit a marked increase in energy transfer during the opening versus closing phases. The energy during opening increased from the midline to 2*y/w*_g_, = 0.21 because of the increased intraglottal pressure at this same location, as observed in Figure 7(a-d). It is believed that this anterior-posterior variation in the intraglottal pressure was due to the three-dimensional flow behavior that occurred. During closure, the energy transfer decreased and became slightly negative at 2*y/w*_g_, = 0.42 because the glottal profile was less divergent at this location. Consequently, the minimum gauge pressure remained positive. The energy transfer, computed as the product of the pressure and the negative surface velocity was, therefore, negative.

The total energy transferred to the VF within the glottis can be computed by assuming a linear variation between the energy at each anterior-posterior location that goes to zero at the anterior-posterior edge, and integrating the energy over the anterior-posterior direction. Note that because the pressure field was assumed symmetric about the anterior-posterior midline, the energy was only computed over half of the VF width, and then multiplied by

2. The aerodynamic energy transfer, *W*, during the opening and closing phases, as well as the total amount, were found to be

*W*_Opening_, = 17.52*µJ, W*_Closure_, = 8.29*µJ, W*_Total_, = 25.81*µJ*, respectively.

The net energy transfer was positive during both opening and closing, which is in contrast with prior numerical^3^ and driven VF model^16^ observations, although only the intraglottal energy transfer was considered in the current work. Nevertheless, these findings are consistent with the theory that the net energy transfer must be positive to ensure self-oscillation^5^. The energy transfer per unit width at the anterior-posterior midline accounted for ≈ 45% of the total energy transfer to the VFs, with the middle third of the glottis accounting for over 80% of the total aerodynamic energy transfer. While prior work^56^ has highlighted how the surface pressure is influenced by anterior-posterior variations, the energy transfer is found to be even more centrally concentrated along the glottal midline due to the multiplicative relationship between the decreased pressure and surface velocity at the anterior-posterior endpoints. The concentration of the energy transfer in the middle of the VF also suggests that more energy is also dissipated along the midline, which coincides with where VF fatigue and damage are most likely to occur^75,76^. This may help explain the prevalence of some VF pathologies such as polyps and nodules in the mid anterior-posterior direction of the VFs^77^.

The variation in energy transfer due to the inferior-superior VF limits over which the aerodynamic pressure is considered has implications for reduced-order VF models as well since, historically, they only consider the contribution to intraglottal energy transfer from medial-lateral force and velocity components^78^, as in the current work. Discrepancies due to the approximation of the medial surface velocity in the current investigations are also likely to influence the energy exchange.

It is interesting to note that the aerodynamic power per unit width (see Figure 9(b)), when multiplied by the total glottal with of 17.0 mm yields a maximum value of ∼ *O*(0.01*W*), which is consistent with prior lumped-element modeling efforts^53,74^. Note that the work of Zan∼artu et al.^74^ reports a similar value for the power transfer as shown in Figure 5 in the article, although the associated text reports an incorrect value due to a typographical error. However, the value of the aerodynamic power in the analogous numerical^3^ and driven VF work^16^ is reported to be two orders of magnitude larger. This difference arises due to computing the power over the entire VF surface, as opposed to just within the glottis. In addition, there also appears to be an inconsistency in how the power was computed in these studies. Namely, the aerodynamic power can be calculated as the product of the pressure integrated over the area and the surface normal velocity, as specified by Equation 5. However, as shown in Figure 14 in ^3^, the aerodynamic pressure at time *t* = 0.057 s can be conservatively estimated to be ≈ 3, 000 Pa over the entire VF surface, which is given to be 1.7 cm in the anterior-posterior direction, and from the same figure, can be estimated to be ≈ 1.5 cm in the inferior-superior direction. The product of these values yields a normal force of ≈ 0.8 N. The surface velocity can be estimated from the slope of the change in orifice width in time, given in Figure 12(a) in^3^. At time *t* = 0.057 s, the slope of the orifice width curve is ≈ 0.7 m*/*s. The resultant power should then be ∼ *O*(0.1*W*). Although this value is larger than those reported in the current physical models, this is likely due to the simple estimation of a constant pressure being distributed over the entire VF surface area for ease of calculation in the current discussion. This clearly overestimates the power transfer. Nevertheless, despite this assumption, the estimated value is still over an order of magnitude smaller than the aerodynamic power, reported as ≈ 8.5 J*/*s at *t* = 0.057 s in Figure 12(b) of Thomson et al^3^. In the same way, it can be inferred that the power magnitude computed in the driven model^16^ is at least an order of magnitude higher than expected.

## IV. CONCLUSIONS

Investigation of the intraglottal aerodynamic pressure was performed with a synthetic self-oscillating VF model in a hemilaryngeal orientation. The key oscillation features of the model such as frequency, open quotient, speed quotient, and glottal distance, were found to have physiologically-relevant values. Therefore, it was deemed a suitable surrogate for investigating the intraglottal pressure distribution.

The intraglottal pressure distribution was found to be highly dependent on unsteady and three-dimensional effects. Unsteady flow effects were reflected in the intraglottal gauge pressure becoming negative during the later stages of VF opening, a feature not captured by prior static VF, steady-flow investigations. This influenced the energy exchange process, decreasing the total amount of energy transferred from the fluid to the VF during the opening phase.

The net aerodynamic energy transfer of the fluid to the structure was also positive during the closing phase, although it was less positive than during opening, thereby satisfying the theory of energy exchange necessary for successful VF oscillation^5^. Immediately preceding VF closure, the energy exchange decreased precipitously as the intraglottal pressure increased due to viscous losses through the narrowing glottal aperture.

Significant variations in the intraglottal aerodynamic pressure were observed between the midline, and the anterior locations along the VF surface. Similarly, the aerodynamic energy transfer varied in the anterior-posterior direction with over 80% of the aerodynamic energy transfer occurring over the middle third of the VF surface. This supports the idea that VF damage occurs along the middle third of the VF length due to the increased dissipation of energy within this region.

Future work aims to use a similar approach to that employed herein to measure intraglottal pressure distributions during the collision phase of the VF in order to improve estimations of collision dose, thereby better predicting VF damage.

## Supporting information

The progression of the intraglottal pressure distribution

## Data Availability

The data that support the findings of this study are available from the corresponding author, upon reasonable request.

## ACKNOWLEDGMENTS

This research was supported by the National Institutes of Health (NIH) National Institute on Deafness and other Communication Disorders Grant P50 DC015446. The content is solely the responsibility of the authors and does not necessarily represent the official views of the National Institutes of Health.

